# Assessment of serological assays for identifying high titer convalescent plasma

**DOI:** 10.1101/2021.03.26.21254427

**Authors:** Christopher W. Farnsworth, James Brett Case, Karl Hock, Rita E. Chen, Jane A. O’Halloran, Rachel Presti, Charles W. Goss, Adriana M. Rauseo, Ali Ellebedy, Elitza S. Theel, Michael S. Diamond, Jeffrey P. Henderson

## Abstract

The COVID-19 pandemic has been accompanied by the largest mobilization of therapeutic convalescent plasma (CCP) in over a century. Initial identification of high titer units was based on dose-response data using the Ortho VITROS IgG assay. The proliferation of SARS-CoV-2 serological assays and non-uniform application has led to uncertainty about their interrelationships. The purpose of this study was to establish correlations and analogous cutoffs between commercially available serological tests (Ortho, Abbott, Roche), a spike ELISA, and a virus neutralization assay using convalescent plasma from a cohort of 79 donors from April 2020. Relationships relative to FDA-approved cutoffs under the CCP EUA were identified by linear regression and receiver operator characteristic curves. Relative to the Ortho VITROS assay, the r^2^ of the Abbott, Roche, the anti-Spike ELISA and the neutralizing assay were 0.58, 0.5, 0.82, and 0.44, respectively. The best correlative index for establishing high-titer units was 3.82 S/C for the Abbott, 10.89 COI for the Roche, 1:1,202 for the anti-Spike ELISA, and 1:200 by the neutralization assay. The overall agreement using derived cutoffs compared to the CCP EUA Ortho VITROS cutoff of 9.5 was 92.4% for Abbott, 84.8% for Roche, 87.3% for the anti-S ELISA and 78.5% for the neutralization assay. Assays based on antibodies against the nucleoprotein (Roche, Abbott) and neutralizing antibody tests were positively associated with the Ortho assay, although their ability to distinguish FDA high-titer specimens was imperfect. The resulting relationships help reconcile results from the large body of serological data generated during the COVID-19 pandemic.

## INTRODUCTION

COVID-19 convalescent plasma (CCP) has been one of the primary therapies deployed in the COVID-19 pandemic. In this current iteration of a classic therapy, serological assays to quantify antibodies to the severe acute respiratory syndrome coronavirus 2 (SARS-CoV-2) spike (S) protein play a critical role in characterizing human immune responses and identifying CCP donors. Commercial SARS-CoV-2 serological assays have accordingly emerged at a rapid pace. Within the first year of the pandemic, more serological assays were available for SARS-CoV-2 than for any other infectious disease, with over 65 emergency use authorizations (EUA) granted for serological testing alone (1). The CDC and Infectious Diseases Society of America (IDSA) have both defined relatively narrow and limited clinical applications for SARS-CoV-2 serological to include CCP donors identification, infection diagnosis in patients more than 14 days from symptom onset, and establishing seroprevalence in populations (2–4). However, the clinical utility of these assays has been questioned (5, 6), in part, due to the challenge of reconciling results from serological assays with clinical outcomes (7–9) and poor agreement between commercial serological assays and virus neutralization assays (10–12).

Identification of CCP with antibody content sufficient for therapeutic use of CCP has emerged as a key quantitative application for SARS-CoV-2 serological assays (2, 5). Anti-S IgG responses in particular were identified early as key correlates of SARS-CoV-2 immunity. At the pandemic’s outset, the absence of an FDA-approved serological assay was a major obstacle to identifying CCP units with sufficient antibody content. A highly sensitive and specific laboratory developed S-based ELISA was quickly developed (13) and used to identify CCP donors with antibodies to SARS-CoV-2 following RT-PCR confirmed infection (14). The initial FDA recommendation was to use a minimum titer of 1:160, with an ideal titer ≥ 1:320, as a criterion for CCP donation (14). A subsequent study demonstrated that high-titer CCP, which was defined as a signal of ≥ 18.45 on the Ortho-Clinical Diagnostics VITROS IgG assay, was associated with lower risk of mortality than those receiving low-titer units in a large retrospective analysis of patients treated through an FDA expanded access protocol (15). A subsequent analysis of this cohort through the Biomedical Advanced Research and Development Authority (BARDA) found that patients receiving CCP with a neutralizing antibody titer > 1:250 experienced lower mortality than those receiving units with titers < 1:250 (16, 17). Neutralizing antibody assays, however, are highly laborious and require biosafety level 3 facilities if using live SARS-CoV-2, limiting their use primarily to research laboratories. As a result, neutralizing assays were correlated with the Ortho Clinical IgG assay, with a minimum signal of 12.0 distinguishing units with high neutralizing titers (18).

In February 2021, the FDA reissued a letter of authorization for CCP with several revisions to the previous EUA (19). Importantly, this included a decision to release only high-titer CCP units for patient use. Cutoffs were provided so that multiple serological assays could be used to define high-titer CCP and previously established titers approved by the FDA were modified. The titer to establish high-titer units with the Ortho Clinical assay was lowered from 12.0 to ≥ 9.5 S/C, and the original anti-S ELISA threshold was raised from 1:320 to ≥ 1:2,880 in an ELISA performed at Mt. Sinai Hospital. The revised EUA also established cutoffs for distinguishing high-titer units using seven other commercial serological assays. For example, the Abbott SARS-CoV-2 IgG assay and the Roche Elecsys anti-SARS-CoV-2 assay were approved for qualifying high-titer units with results ≥ 4.5 signal-to-cutoff (S/C) and ≥ 109 cutoff index (COI), respectively.

Little published literature is available to correlate neutralizing antibody titers, commercial serological assays, and anti-S ELISAs. Several studies have assessed the positive percent agreement and negative percent agreement (PPA and NPA) between assays (10, 11, 20). However, the signal from other commercial, serological assays that best correlate to anti-S ELISA titers of 1:320, neutralizing titers of 1:250, and the Ortho Clinical S/C of 12.0, have not been determined. The purpose of our study was to establish correlations and analogous cutoffs between widely used commercial serological assays, anti-S ELISA, and neutralizing assays with authentic SARS-CoV-2. The resulting relationships will help reconcile results from the large body of serological studies and CCP trials results that continue to emerge during the COVID-19 pandemic.

## MATERIALS AND METHODS

### Human subjects

This study was approved by the Washington University Institutional Review Board. Serum specimens were drawn on patients with RT-PCR-confirmed SARS-CoV-2 infection at least 14 days after infection and prior to donation of convalescent plasma. Patient reported demographic information including age, gender, race, comorbidities, and duration of symptoms were collected by survey on each patient. After collection, specimens were immediately frozen in 100 µL aliquots and stored at -80°C until further analysis.

### Assays

Specimens were thawed at room temperature and analyzed within 3 days. Three commercial serological assays and an anti-S ELISA granted EUA at Mt. Sinai Hospital, but used on a research basis for this study, were used to directly measure antibody levels in serum specimens. These assays detected antibodies to SARS-CoV-2 S or nucleocapsid proteins. The anti-S ELISA was performed as previously described (13). In short, plasma specimens were diluted to 1:30 in PBS, then serially diluted to 1:65,610 in a 96-well plate (Corning). Wells were washed, incubated with a secondary anti-human IgG, followed by another wash step. Wells were then incubated with o-Phenylenediamine dihydrochloride (Sigma-Aldrich) followed by a stop solution (3M hydrochloric acid). The optical density was then measured at 490 nm and the cutoff for a positive result was determined as an optical density that was three standard deviations above the mean signal from a negative control specimen run with each plate. This signal was extrapolated from the generated curves to quantify the titer (21).

An authentic SARS-CoV-2 neutralization assay was used to measure neutralizing antibody titers. Focus reduction neutralization assays were performed as previously described (10). SARS-CoV-2 strain n-CoV/USA_WA1/2020 was obtained from the Centers for Disease Control. Virus was propagated in Vero E6 cells in Dulbecco’s Modified Eagle Medium (DMEM, Corning) that was supplemented with 10% FBS, glucose, L-glutamine, and sodium pyruvate. Patient sera were diluted and incubated with 1×10^2^ focus forming units (FFU) of SARS-CoV-2 for 1 h at 37°C. The plasma/virus complex was then added to Vero E6 monolayers at 37°C for 1 h. After overlaying with methylcellulose, cells were harvested at 30 h, methylcellulose was removed, and fixed with 4% paraformaldehyde (PFA) for 20 min. Plates were washed and incubated with 1 µg/mL anti-S antibody (CR3022) and HRP conjugated goat anti-human IgG. Infected cells were visualized with TrueBlue peroxidase substrate (KPL) and quantified using an ImmunoSpot microanalyzer (Cellular Technologies). A minimum of eight dilutions was performed for each specimen, a standard curve generated, and the 1/log10 plasma dilution (EC50) determined as the dilution at which 50% of the cells were infected.

All specimens were analyzed on three commercially available serological assays. The Ortho Clinical VITROS SARS-CoV-2 IgG assay was performed on an Ortho Clinical VITROS 5600 Immunodiagnostic System and targets antibodies to the S protein. The Abbott SARS-CoV-2 IgG assay was performed on an Abbott Architect i2000 and detects antibodies to the nucleocapsid protein. The Roche Elecsys anti-SARS-CoV-2 assay was performed on a Cobas e601 and identifies antibodies to the nucleocapsid protein. All commercial assays have FDA EUA as qualitative methods and were performed and interpreted according to the manufacturer’s instructions. The positive cutoffs for each assay are 1.0 (S/C), 1.4 (S/C), and 1.0 (COI) for the Ortho Clinical, the Abbott, and the Roche assays, respectively. All three assays report a numeric signal to cut-off that is the amount of signal generated by the sample for each assay relative to the signal from a single calibrator.

### Statistical analysis

Association between assays were compared with least squares regression to calculate intercept, slope and r^2^. Ideal cutoffs from linear regression were established by interpolating relative to each cutoff. Receiver operator curves (ROC) were also plotted to assess the ideal cutoffs using Youden Index to establish cutoff with maximum positive and negative percent agreement. Final cutoffs for distinguishing high- and low-titers units by each assay were established by averaging across all cutoffs.

## RESULTS

### COVID-19 convalescent plasma donors

Serum specimens were obtained from 79 adults at Washington University/Barnes-Jewish Hospital Medical Center in St. Louis, Missouri, U.S.A., with a history of positive SARS-CoV-2 RT-PCR testing who expressed interest in donating CCP between 4/6/2020-4/29/2020. The median age was 49 (range 20-69) (**Table 1**). 55.7% of patients were female, 91.1% were white. The most common comorbidity was asthma. Only 2 patients (2.5%) were hospitalized, and the median duration of symptoms was 12 days (range; 1-31). The median time from symptom onset to positive RT-PCR result was 4 days (range; 0-20).

**Table 1.** Convalescent plasma donors’ characteristics

| Variable | Total n=79 (%) |
| --- | --- |
| Age (median [range]) | 49 (20-69) |
| Sex |  |
| Female | 44 (55.7) |
| Male | 35 (44.3) |
| Race |  |
| White | 72 (91.1) |
| Black | 4 (5.1) |
| Asian | 2 (2.5) |
| Other | 1 (1.3) |
| Comorbidities |  |
| Asthma | 15 (19) |
| Lung disease | 0 (0) |
| Heart disease | 2 (2.5) |
| Hypertension | 13 (16.5) |
| Diabetes mellitus | 3 (3.8) |
| Cancer | 6 (7.6) |
| Autoimmune disease | 5 (6.3) |
| Other | 28 (35.4) |
| Hospitalization | 2 (2.5) |
| Duration of symptoms in days (median [range]) | 12 (1-31) |
| Days from symptom onset to positive test (median [range]) | 4 (0-20) |

Anti-S ELISA IgG titers in this cohort spanned four orders of magnitude (1:21 – 1:17,278) with a median titer of 1:2,315 (**Fig 1A)**. A broad range of responses also was evident among commercial serological assays. These results are consistent with substantial variability in antibody responses to SARS-CoV-2 proteins among recovered adults. The median signal of the Ortho Clinical assay was 15.4 S/C (95% CI: 12.7-18.0 S/C) (**Fig 1B**), the Abbott assay was 5.2 S/C (95% CI: 4.3-6.1 S/C) (**Fig 1C**), and the Roche assay was 23.94 COI (95% CI: 13.8-37.1 COI) (**Fig 1D**). As with the anti-S ELISA IgG, live virus neutralization titers spanned a broad range (1:20 – 1:3,622), with a median titer of 1:316 (95% CI: 1:251-1:398) (**Fig 1E**). These results are consistent with substantial variability in neutralizing antibody responses to SARS-CoV-2 proteins among recovered adults (12).

**Figure 1.**
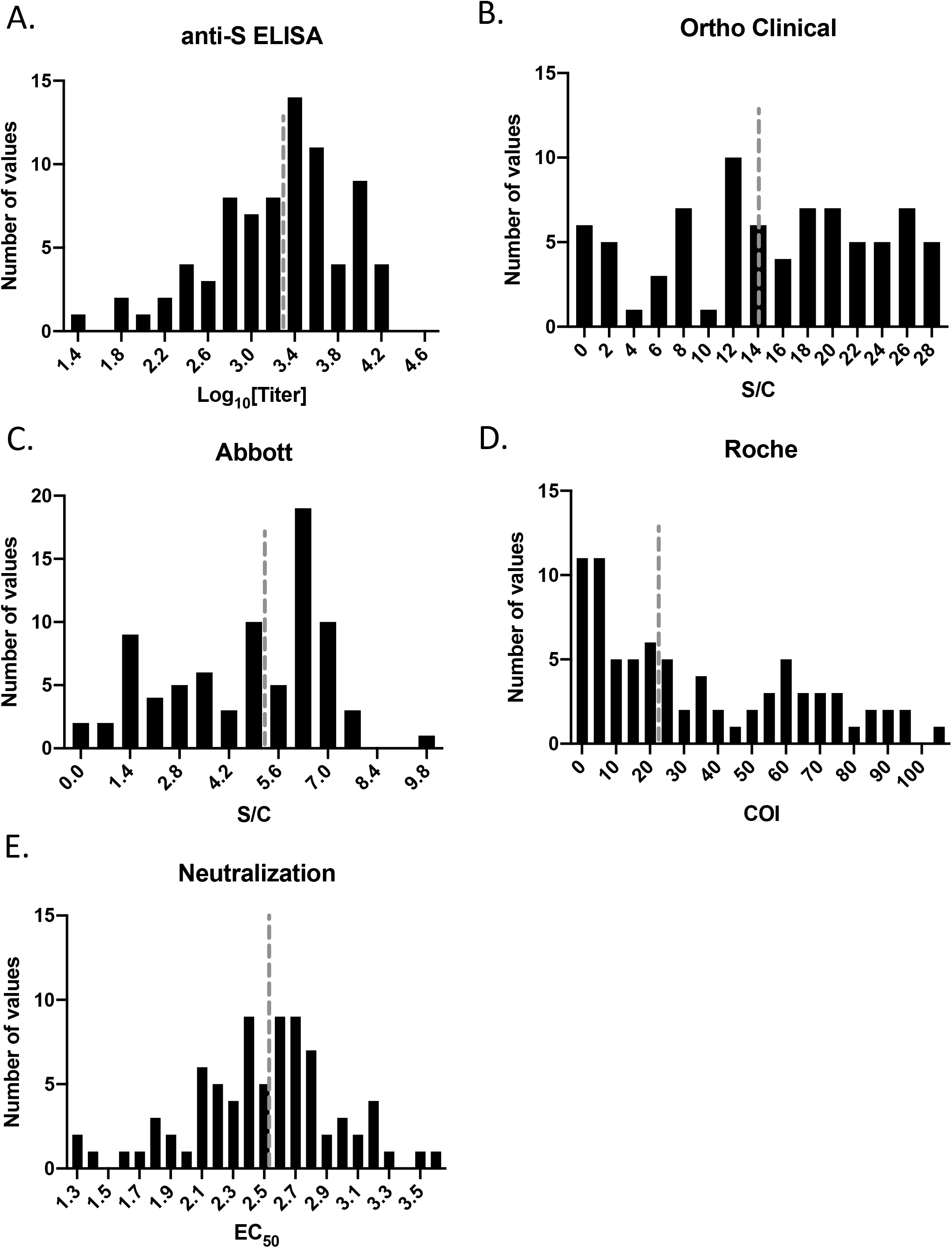
Histogram of each assay for 79 convalescent plasma donors with confirmed SARS-CoV-2 infection. Dashed line is the median.

### Serological characteristics of donors

Linear relationships between each commercial assay were defined relative to the anti-S ELISA IgG titer, Ortho-Clinical assay, and neutralization titer (**Supplemental Figure 1**). Slopes and intercepts for each commercial assay relative to the anti-S ELISA, Ortho Clinical IgG, and neutralization assay are found in **Table 2**. For the anti-S ELISA IgG titer, the initial FDA titer of 1:320 was used to establish high and low titers (**Supplemental Fig. 1A**). Interpolated signal by the Ortho Clinical was 5.13 S/C (4.12-6.04), for the Abbott was 1.39 S/C (0.59 to 1.91) and for the neutralization assay was 1.82 (1.68 to 1.93). Due to poor fit, an interpolated cutoff could not be calculated for the Roche assay. For the Ortho Clinical assay, the cutoff of 12.0 S/C was used to distinguish high- and low-titer units (**Supplemental Fig 1B**). The interpolated signal by the Abbott assay was 3.62 S/C (3.1 to 4.07), for the Roche assay was 7.46 COI (4.78 to 10.73), for the ELISA was 2.96 (2.87 to 3.04), and for the neutralization assay was 2.25 (2.09 to 2.37). Low- and high-titer cutoffs were also calculated at the Ortho Clinical cutoff of 4.62 S/C and 18.45 S/C, respectively. For the neutralization assay, a single cutoff of 1:250 was used to determine low- and high-titer units (**Supplemental Fig 1C**). Relative to the neutralization assay, the interpolated cutoffs for the Ortho Clinical was 12.39 S/C (10.38 to 14.16), for the Abbott was 3.92 S/C (3.14 to 4.57), for the Roche assay was 8.45 COI (3.48 to 15.89), and for the anti-S IgG ELISA was 3.05 (2.91 to 3.18).

**Table 2.** Linear fit and interpolated cutoffs for each assay

|  |  | Ortho Clinical | Abbott | Roche | anti-S ELISA | Neutralizing |
| --- | --- | --- | --- | --- | --- | --- |
| anti-S ELISA Titer | Intercept | -632.5 | -359.5 | 467.9 |  | -2875 |
|  | (95% CI) | (-1062 to -203.1) | (-993.0 to 210.2) | (53.7 to 882.2) |  | (-4281.0 to -1468.0) |
|  | Slope (95% CI) | 121.2 (96.05 to 146.4) | 337.3 (221.7 to 452.9) | 21.0 (11.8 to 30.2) |  | 1625.0 (1070.0 to 2180.0) |
|  | r<sup>2</sup> | 0.54 | 0.3 | 0.21 |  | 0.31 |
|  | FDA cutoff 1:320 (95% CI) | 5.13 (4.12-6.04) | 1.39 (0.59 to 1.91) | NA* |  | 1.82 (1.68 to 1.93) |
|  | FDA cutoff 1:2,884 | 17.75 (16.8 to 18.75) | 5.64 (5.23 to 6.07) | 52.53 (43.05 to 65.31) |  | 2.69 (2.62 to 2.774) |
| Ortho-Clinical |  |  |  |  |  |  |
|  | Intercept (95% CI) |  | 1.72 (-1.1 to 4.55) | 9.64 (7.29 to 11.99) | -24.549 (-28.74 to -20.24) | -14.77 (-22.44 to -7.1) |
|  | Slope (95% CI) |  | 2.83 (2.29 to 3.38) | 0.16 (0.1 to 0.21) | 12.12 (10.83 to 13.4) | 11.91 (8.89 to 14.94) |
|  | r<sup>2</sup> |  | 0.58 | 0.5 | 0.82 | 0.44 |
|  | FDA cutoff 12 (95% CI) |  | 3.62 (3.1 to 4.07) | 7.46 (4.78 to 10.73) | 3.01 (2.87 to 3.04) | 2.25 (2.09 to 2.37) |
|  | High Cutoff 18.45(95% CI) |  | 5.91 (5.45 to 6.46) | 41.02 (2.73 to 84.33) | 3.54 (3.46 to 3.64) | 2.79 (2.66 to 2.96) |
|  | Low Cutoff 4.62 (95% CI) |  | 1.02 (0.03 to 1.73) | 1.06 (0.43 to 1.93) | 2.4 (2.28 to 2.51) | 1.63 (1.3 to 1.83) |
| Neutralizing |  |  |  |  |  |  |
| (EC50) | Intercept (95% CI) | 1.94<br>(1.77 to 2.1) | 9.64<br>(7.29 to 11.99) | 2.09<br>(1.9 to 2.28) | 0.59<br>(0.24 to 0.94) |  |
|  | Slope (95% CI) | 0.04<br>(0.03 to 0.05) | 0.16<br>(0.1 to 0.21) | 0.34<br>(0.2 to 0.47) | 0.59<br>(0.48 to 0.69) |  |
|  | r<sup>2</sup> | 0.44 | 0.5 | 0.24 | 0.61 |  |
|  | FDA cutoff 250 (95% CI) | 12.39<br>(10.38 to 14.16) | 3.92<br>(3.14 to 4.57) | 8.45<br>(3.48 to 15.89) | 3.09<br>(2.97 to 3.2) |  |
|  | High Cutoff 500 (95% CI) | 20.49<br>(18.56 to 22.94) | 6.28<br>(5.58 to 7.23) | 67.04<br>(33.4 to > assay) | 3.61<br>(3.49 to 3.75) |  |
|  | Low Cutoff 125 (95% CI) | 12.39<br>(10.38 to 14.16) | 1.58<br>(0.11 to 2.48) | 1.07<br>(0.01 to 2.78) | 2.58<br>(2.38 to 2.72) |  |

ROC curves were generated for each serological assay using the Ortho Clinical cutoff of 12 S/C (**Fig 2A**) and the neutralizing cutoff of 1:250 (**Fig 2B**). Relative to the Ortho Clinical assays, all assays had an AUC > 0.8, with the anti-S IgG ELISA having the best correlation (AUC = 0.927). The assay with the greatest correlation with the neutralizing assay was the anti-S IgG ELISA, with an AUC of 0.856 (0.771-0.941). Final cutoffs were established by averaging the ideal cutoffs from linear regression in **Table 2** and the ideal cutoffs from the ROC curves using Youden’s Index. Using this approach, the average cutoff for distinguishing high- and low-titer units by the Abbot assay was 3.82 S/C, the Roche assay was 10.89 COI, the anti-S IgG ELISA was 3.08, 1:200 for the neutralization assay and 14.14 S/C for the Ortho-Clinical assay (**Table 3**). ROC curves were also generated relative to the low and high Ortho-Clinical cutoffs of 4.62 S/C and 18.45 S/C, respectively (**Supplemental Fig 2**) and for the low and high neutralizing titers of 1:150 and 1:500, respectively (**Supplemental Fig 3**).

**Table 3.**
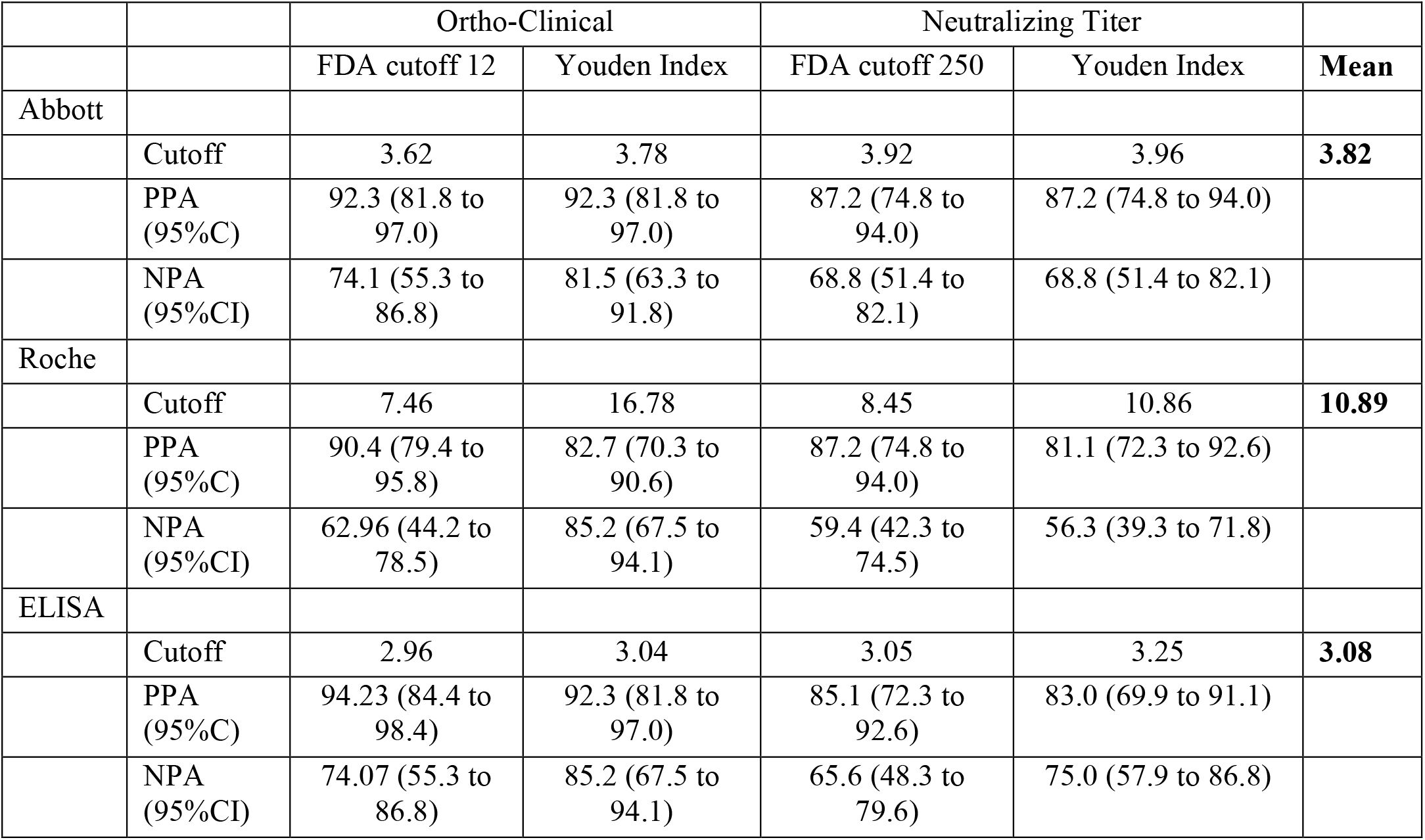
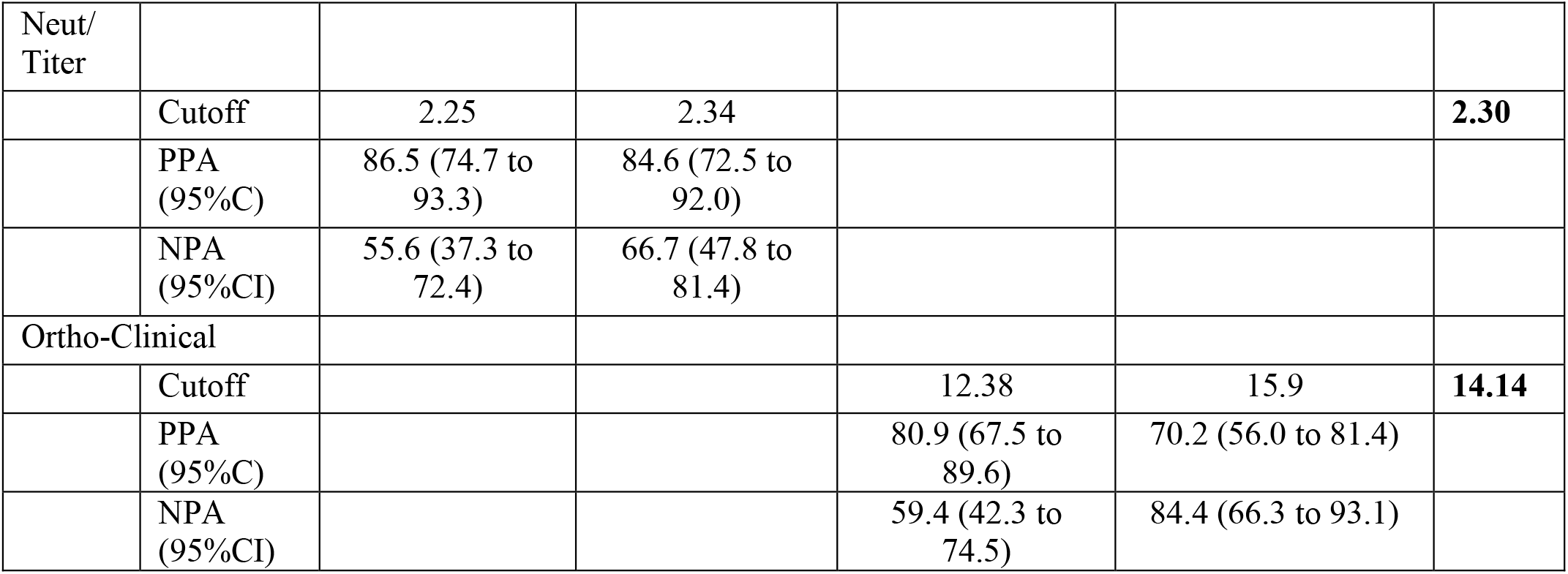
PPA and NPA for each assay at various cutoffs relative to the Ortho Clinical assay and Neutralizing titers

**Figure 2.**
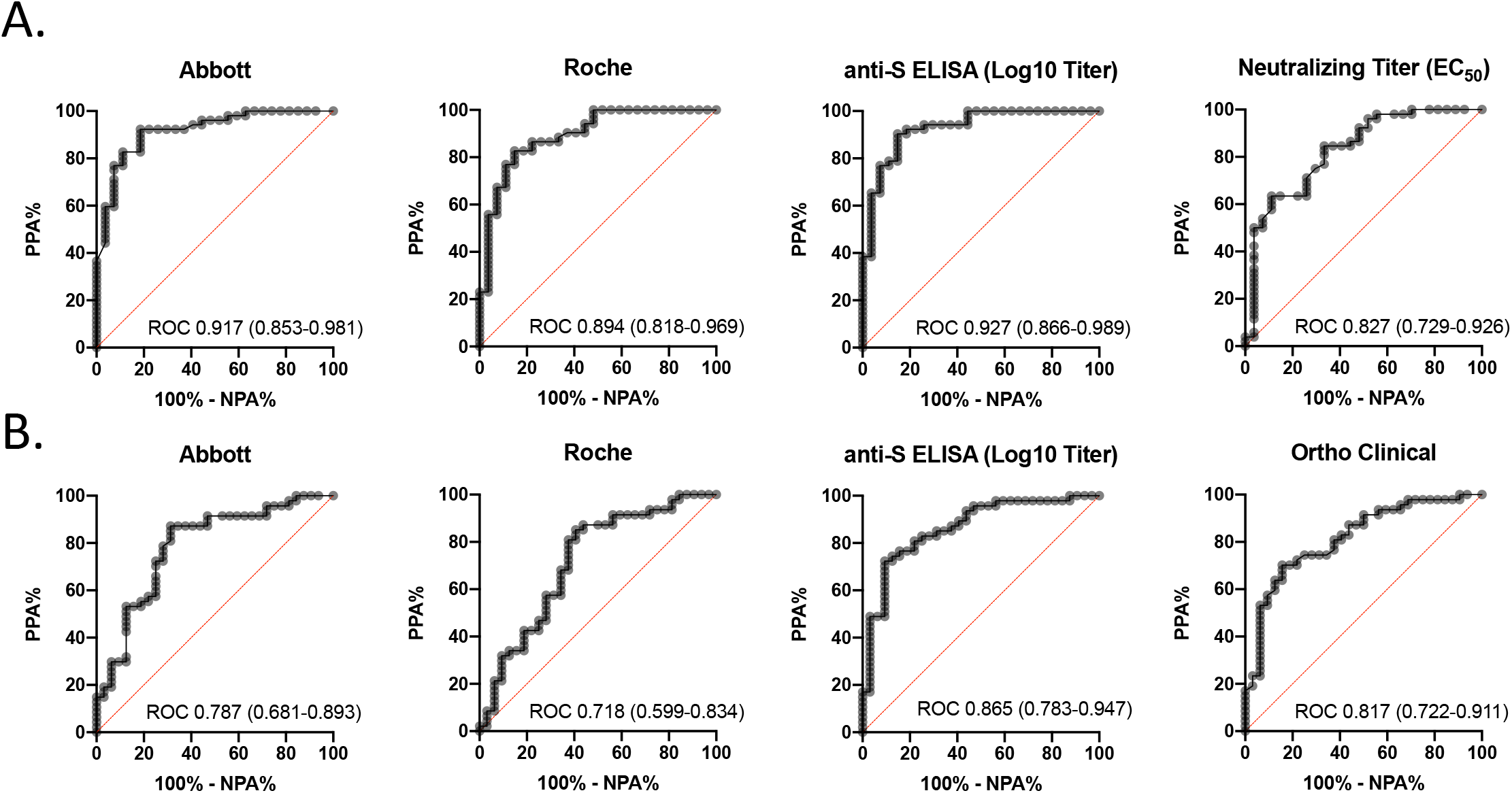
ROC Curves for serological SARS-CoV-2 assays with **A**. the Ortho-Clinical IgG assay using a cutoff of 12 to distinguish high titers and **B**. neutralizing assay using a cutoff of 250.

Specimens were segregated as low- or high-titer using the Ortho Clinical cutoff of 9.5 S/C or 12 S/C and scatterplots generated (**Fig. 3A and B**). Using the cutoffs established in **Table 3** (dotted black lines), all four assays (Abbott, Roche, ELISA, and neutralization) demonstrated comparable performance relative to the Ortho Clinical cutoffs of 9.5 S/C and 12 S/C for identifying patients with high and low antibody titers. Decreasing the signal for identifying high-titer plasma on the Ortho Clinical assay led to improved NPA and PPA with the Abbott and Roche assay and an improved NPA with a modest decrease in PPA with the anti-S IgG ELISA and the neutralization titer (**Supplemental Table 1**). The overall agreement using the derived cutoffs with the Ortho Clinical assay cutoff of 9.5 S/C was 92.4% for Abbott, 84.8% for Roche, 87.3% for the anti-S ELISA and 78.5% for the neutralization assay. Relative to the FDA Abbott cutoff of 4.5 S/C (dashed gray line) for identifying high-titer units, 5 additional specimens would have been labeled as low-titer by the Abbott but high-titer by the Ortho Clinical assay. Using the FDA cutoff of ≥109 COI for the Roche assay, all 79 specimens would have been qualified as low-titer units. Specimens also were segregated as low- or high-titer using the neutralization assay cutoff of 1:250 with similar results (**Fig. 3C**). The tiered Ortho Clinical and neutralizing cutoffs used to identify patients with low medium and high titers are found in **Supplemental Figures 4 and 5**, respectively. Patients with high ratios of nucleocapsid to S as measured by the Abbott and Ortho assays were more likely to have low neutralizing antibody titers (**Supplemental Figure 6)**.

**Figure 3.**
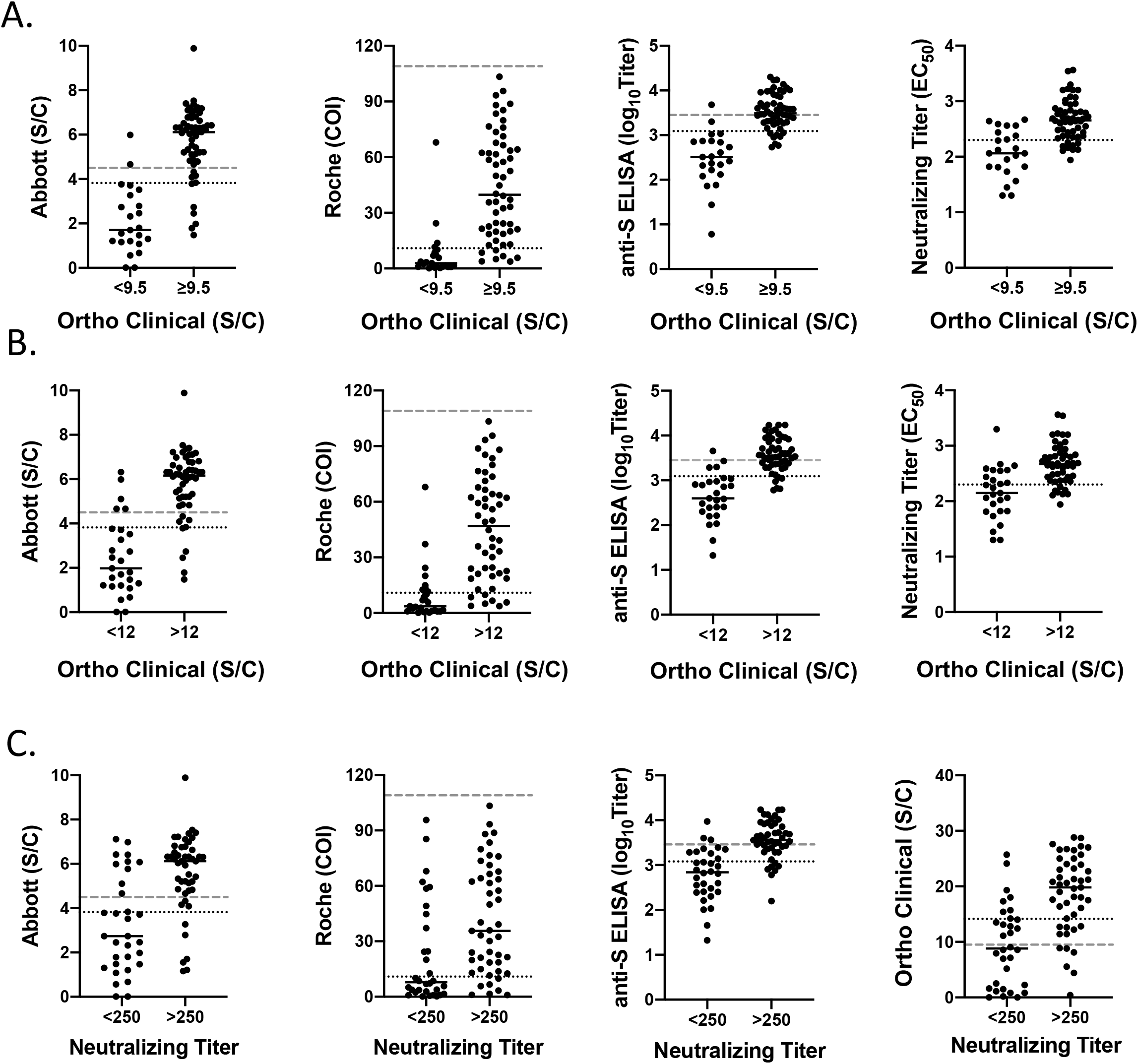
Scatter plots of EUA serological SARS-CoV-2 assays using the ideal cutoffs identified by linear regression and ROC curves relative to **A**. Ortho-Clinical cutoff of 9.5, **B**. Ortho Clinical cutoff of 12.0 and **C**. a neutralizing titer cutoff of 1:250. Dotted line represents the mean cutoffs identified in Table 3. Dashed lines represent the FDA approved cutoffs from EUA.

## DISCUSSION

Despite accumulating evidence of associations between commercial serological assay values and neutralizing antibody titers with human immunity and CCP efficacy, few published studies permit correlation between the assay formats in use. Here, we tested three widely used commercial serological assays, an EUA anti-S IgG ELISA, and neutralizing antibodies and correlated each assay with the ideal cutoffs for establishing high-titer plasma.

An important finding from this study is that commercial assays and the anti-S ELISA performed similarly for identifying specimens with high neutralization titers. Our approach using linear regression for the Ortho Clinical assay with the FDA-established cutoff of 12 S/C and the neutralizing titer of 1:250, coupled with ROC curves that established maximal PPA and NPA-identified cutoffs made these assays largely interchangeable for identifying high-titer CCP. The antigenic target of the assay did not change the PPA and NPA, with assays measuring antibodies to the viral S protein performing similarly to those measuring antibodies to the nucleocapsid protein. This finding is similar to other studies from acutely infected patients with severe symptoms and patients with mild symptoms (10, 22).

It is notable that FDA’s reissued CCP authorization letter incorporated multiple EUA serological assays, several of which are included in this report (16). The cutoff for the Abbott assay established here (≥ 3.82 S/C) is similar to the cutoff of ≥ 4.5 S/C established by the FDA. This is despite the lowered Ortho Clinical assay S/C cutoff (from 12 to 9.5) in the reissued CCP EUA (16). Nonetheless, the Abbott assay cutoff of 3.82 S/C had better PPA with the Ortho Clinical cutoffs of 9.5 S/C and 12.0 S/C, without sacrificing NPA. In contrast, the FDA-approved cutoff of 109 COI for the Roche assay would have disqualified all units as low-titer, with a signal approximately 10-fold higher than the ideal cutoff identified in this study. The derived cutoff from this study with the anti-S IgG ELISA (1:1,202) also was lower than that established by the FDA (1:2,880). This resulted in a considerable difference in PPA, with far more convalescent donor units being excluded under the FDA cutoffs for ELISA than for the Ortho Clinical assay. The cutoff identified in our study that best distinguishes neutralizing titers ≥ 1:250 in the Ortho Clinical assay was 14.14 S/C, higher than the original cutoff of 12.0 S/C from the FDA. An S/C of 9 on the Ortho Clinical assay correlated to a neutralizing titer of ∼1:100. This is notably similar to the neutralizing titer of 1:104 we found to be sufficient to reduce weight loss in mice (23). Nevertheless, the neutralizing assay used in this study cannot be assumed to perform similarly to the assay used in the BARDA study (14, 15) due to non-standardization of SARS-CoV-2 strains, cell lines, and reagents/procedures. These differences underlie the difficulties in harmonizing SARS-CoV-2 serological assay results. These discrepancies should be considered when attempting to use any serological assay as a proxy for measuring neutralizing antibodies.

Several studies demonstrate survival benefits with early, high-titer CCP administration and in patients with hematological malignancies, implying a continued need to identify CCP or immune globulin donors (15, 24–26). This study attempts to harmonize several commercially available assays that have been extensively studied and published. While numerous studies have addressed the analytical performance of available serological assays, little correlative information is available in the published literature to relate multiple serological assay results. Many blood centers in the US are currently using or switching to Ortho Clinical IgG assay for identification of high-titer units. The data presented here suggest that multiple commercial assays could be used to identify CCP donors with high levels of neutralizing antibodies. This study may also provide useful information for contextualizing previous seroprevalence studies and multiple CCP studies across the literature emerging from this pandemic.

There are several limitations associated with this study. Among the greatest limitations is the lack of standardization between assays, even among the same manufacturers. This was previously noted with the neutralization assay, though the same is true among commercial assays. Since several of the assays have been designated as qualitative (*i.e*. the Roche, Abbott, and Ortho Clinical assays), there is limited evidence that semi-quantitative results are comparable between different instruments by the same manufacturer above the cutoff. For example, since there is no material to verify linearity at higher concentrations, a result of 15 S/C at one institution using the Ortho Clinical assay may vary from the Ortho Clinical assay at another institution. This may underlie the differences between the established cutoff and FDA cutoff for the Roche assay. In general, this problem will continue to plague the field until quantitative assays are universally adopted and standardized to SARS-CoV-2 antibody reference material, such as that recently released by the World Health Organization (27). This is further complicated by unclear direction as to how to report a qualitative assay result as quantitative under an EUA, which does not permit modification of the manufacturer’s Instructions for Use. Another limitation of the current study is that a limited number of assays were evaluated, limiting the generalizability of results. It is also important to note that these specimens were obtained early during the course of the pandemic, and that continued viral evolution (which may lead to extensive antigenic changes in the S protein) means that the quantitative relationships in this manuscript could become outdated. Ongoing studies are required to confirm the relationships established here in patients infected with SARS-CoV-2 variants and using neutralizing assays that utilize SARS-CoV-2 variants. Finally, while these results provide evidence of that the cutoffs identified by the FDA are helpful for identifying high-titer CCP units, there were several specimens by each assay that were not in agreement. These specimens demonstrate the requirement for further study before the cutoffs proposed by the FDA are modified.

In conclusion, we demonstrate that assays based on nucleoprotein antibodies (Roche, Abbott) and neutralization were positively associated with Ortho assay results (anti-S), though their ability to distinguish FDA high-titer specimens was marginal. Association with a traditional ELISA serologic test was high. All assays were positively associated with neutralization titers, though associations were strongest with S-based assays.

## Data Availability

Data will be made available upon request at the discretion of the authors

## AKNOWLEDGEMENTS

This study utilized samples obtained from the Washington University School of Medicine’s COVID-19 biorepository, which is supported by: the Barnes-Jewish Hospital Foundation; the Siteman Cancer Center grant P30 CA091842 from the National Cancer Institute of the National Institutes of Health; and the Washington University Institute of Clinical and Translational Sciences grant UL1TR002345 from the National Center for Advancing Translational Sciences of the National Institutes of Health. J.P.H. acknowledges support from National Institute of Diabetes and Digestive and Kidney Diseases grant RO1DK111930 and the Longer Life Foundation. The content is solely the responsibility of the authors and does not necessarily represent the view of the NIH.

